# Emergence of a new SARS-CoV-2 variant from GR clade with a novel S glycoprotein mutation V1230L in West Bengal, India

**DOI:** 10.1101/2021.05.24.21257705

**Authors:** Rakesh Sarkar, Ritubrita Saha, Pratik Mallick, Ranjana Sharma, Amandeep Kaur, Shanta Dutta, Mamta Chawla-Sarkar

## Abstract

India is currently facing the devastating second wave of COVID-19 pandemic resulting in approximately 4000 deaths per day. To control this pandemic continuous mutational surveillance and genomic epidemiology of circulating strains is very important. In this study, we performed mutational analysis of the protein coding genes of SARS-CoV-2 strains (n=2000) collected during January 2021 to March 2021. Our data revealed the emergence of a new variant in West Bengal, India, which is characterized by the presence of 11 co-existing mutations including D614G, P681H and V1230L in S-glycoprotein. This new variant was identified in 70 out of 412 sequences submitted from West Bengal. Interestingly, among these 70 sequences, 16 sequences also harbored E484K in the S glycoprotein. Phylogenetic analysis revealed strains of this new variant emerged from GR clade (B.1.1) and formed a new cluster. We propose to name this variant as GRL or lineage B.1.1/S:V1230L due to the presence of V1230L in S glycoprotein along with GR clade specific mutations. Co-occurrence of P681H, previously observed in UK variant, and E484K, previously observed in South African variant and California variant, demonstrates the convergent evolution of SARS-CoV-2 mutation. V1230L, present within the transmembrane domain of S2 subunit of S glycoprotein, has not yet been reported from any country. Substitution of valine with more hydrophobic amino acid leucine at position 1230 of the transmembrane domain, having role in S protein binding to the viral envelope, could strengthen the interaction of S protein with the viral envelope and also increase the deposition of S protein to the viral envelope, and thus positively regulate virus infection. P618H and E484K mutation have already been demonstrated in favor of increased infectivity and immune invasion respectively. Therefore, the new variant having G614G, P618H, P1230L and E484K is expected to have better infectivity, transmissibility and immune invasion characteristics, which may pose additional threat along with B.1.617 in the ongoing COVID-19 pandemic in India.

## Introduction

Since the emergence of SARS-CoV-2, which is the cause of COVID-19 pandemic, in December 2019, the virus has continuously been evolving by acquiring mutations throughout the genome with the progression of the pandemic [1]. Continuous efforts are being made towards SARS-CoV-2 genome sequencing and genomic epidemiology to investigate the virus evolution, their pathogenic potential and transmission. Gradual accumulation of mutation within the genome resulted in the delineation of circulating SARS-Cov-2 strains into different clades (L, S, O, V, G, GH, GR, GV, and GRY according to GISAID nomenclature) clades, lineages and sublineages [2]. Special attention is given to mutations which have evolved within the S glycoprotein due to their role in host cell receptor binding and entry, immune invasion and antibody neutralization [3]. Not surprisingly, most of the new mutations have been observed within the S glycoprotein, which could provide fitness advantage in terms of enhanced infectivity, transmissibility, immune invasion or antibody neutralization.

D614G, the first mutation appeared within the S glycoprotein has been shown to be responsible for better infectivity and transmissibility of virus [4-6]. This mutation is the characteristic of most circulating clades (G, GH, GR, GV, GRY) and 4 newly emerged Variant of Concerns (VOCs) UK variant or B.1.1.7 (evolved within the GR clade), South African variant or B.1.351 (evolved within the GH clade), Brazilian variant or P.1 (evolved within the GR clade) and California variant or B.1.427/B.1.429 (evolved within the GH clade). Along with the clade specific mutations, these variants have their own set of characteristics mutations including several mutations in the S glycoprotein like Δ69/70, Δ147, N501y, A570D, P681H, T716I, S982A, D1118H in UK variant; D80A, D251G, Δ241/242/243, K417N, E484K, N501Y, A701V in South African variant; L18F, T20N, P26S, D138Y, R190S, K417T, E484K, N501Y, H655Y, T1027I in Brazilian variant; and S13I, W152C, L452Q in California variant [7-12].

Recently, India has witnessed the emergence of two new variants namely B.1.617, first detected in Maharashtra on 5^th^ October 2020, and B.1.618, first detected in West Bengal on 25^th^ October. B.1.617 is characterized by the S glycoprotein mutations L452R, E484Q, D614G, and P681R [13-15]. It has three sublineages B.1.617.1 (S protein mutations: T95I, G142D, E154K, L452R, E484Q, D614G, P681R and Q1071H), B.1.617.2 (S protein mutations: T19R, G142D, Δ156/157, R158G, L452R, T478K, D614G, P681R, D950N) and B.1.617.3 (S glycoprotein mutations: T19R, G142D, L452R, E484Q, D614G, P681R and D950N) (https://www.cdc.gov). Among these three sublineages, B.1.617.3 was first detected in October, 2020. However, its prevalence remained very low compared to the other two sublineages, both of which were first identified in December, 2020. Frequency of B.1.617 (all sublineages) started to rise significantly in February 2021, resulting in devastating second wave of COVID-19 pandemic in India [13]. Spread of this variant outside India was first reported from UK, USA and Singapore in late February 2021. By 13 May 2020, this variant has been detected in about 60 countries with more than 4500 confirmed sequences. On 7 May 2020, Public Health Authorities of England declared B.1.617.2 as VOC. On 11 May, WHO also declared this variant as VOC under the name VOC-21ARP-02 based on evidences that this variant is at least as transmissible as UK variant and less sensitive to antibody neutralization. The second Indian variant, B.1.618, also known as triple mutant, has four mutations ΔH146, ΔY147, E484K and D614G in the S glycoprotein. Members of this lineage have also been found to spread in other countries like US, Singapore, Switzerland and Finland. The prevalence of this mutant has been growing significantly in West Bengal, but remains unidentified in other states of India [15].

In the current scenario, tracking mutations within these newly emerged variants and also investigating the emergence of new variants is crucial to take necessary preventive measures for controlling the COVID-19 pandemic in India. With the aim of tracing new mutations within the circulating SAR-CoV-2 strains in India during January 2021 to February 2021, we have identified the emergence of a new variant in West Bengal, India. This new variant is characterized by the presence of 12 co-existing mutations including E484K, D614G, P681H and V1230L in S glycoprotein.

## Materials and Methods

### Sequence retrieval

High coverage full genome sequences of 2000 SARS-CoV-2 strains, collected during January 2021 to March 2021 from India, were retrieved from Global GISAID database on 15 May 2021 [16]. The genome sequence of the prototype SARS-CoV-2 strain hCoV-19/Wuhan/WIV04/2019 (GISAID accession no. EPI_ISL_402124) and several clades/variants was also downloaded from GISAID database.

### Screening of mutations

In this study we have performed non-synonymous mutational analysis of 25 proteins (NSP1-NSP16, S glycoprotein, NS3, E, M, NS6, NS7a, NS7b, NS8 and N) encoded by each of the 2000 SARS-CoV-2 genome sequences submitted from India. For performing mutational analysis, each of the 25 protein coding regions of circulating SARS-CoV-2 genomes as well as prototype genome (hCoV-19/Wuhan/WIV04/2019) was cut and subsequently translated to amino acid sequence by using TRANSEQ nucleotide-to-protein sequence conversion tool (EMBL-EBI, Cambridgeshire, UK). Next, each of the 25 protein sequences of 2000 SARS-CoV-2 genomes was aligned with the corresponding protein sequence of the prototype strain by using MEGA software (Version X) and observed for amino acid substitutions [17].

### Phylogenetic analysis

A phylogenetic dendrogram was constructed based on the whole genome sequences of 111 SARS-CoV-2 strains, including 38 sequences of the new variant and 73 reference sequences of different clades/variants (11 reference sequences of Indian variant B.1.617; 5 reference sequences of each of G clade, GR clade, GV clade, L clade, V clade, GRY clade/UK variant, South African variant, Brazilian variant, California variant, Nigerian variant and Indian variant B.1.618; 4 reference sequences of S clade; 3 reference sequences of GH clade), using Molecular Evolutionary Genetics Analysis (MEGA) version X [18]. Initially, genome sequences of 122 SARS-CoV-2 strains were aligned multiple alignment program MAFT version 7 [19]. The alignment file was then used to build phylogenetic tree by maximum-likelihood method using general time reversal (GTR) statistical model with 1000 bootstrap replicates [17]. We further performed the phylogenetic analysis of the 70 sequences of the new variant with the sequence analysis web app Nextclade^beta^ version 0.14.3 (https://clades.nextstrain.org/).

### Results and Discussion

By performing the whole genome mutational analysis of 2000 SARS-CoV-2 strains collected during January 2021 to March 2021 from India, we have identified 70 SARS-CoV-2 strains having new set of 11 co-existing mutations: NSP4 has two mutations D279N and L353F; NSP8 has a single mutation of V26F; NSP12 has a single mutation of P323L; S gene is characterized by three mutations D614G, P681H and V1230L; NS3 has a single mutation of G172C; NS8 has a single mutation of V62L; and N gene is characterized by two mutations R203K and G204R **(Table 1, Supplementary Table 1)**. Interestingly, among 70 sequences, 16 sequences also harbored E484K mutation along with 614G, P681H and V1230L in the S glycoprotein. This new variant has evolved within the GR clade (Pango lineage B.1.1 or 20B), which is characterized by four co-existing mutations D614G in S glycoprotein, P323L in NSP12, and R203K and G204R in N protein, all of which are present within this new variant. Phylogenetic analysis of 38 representative strains of this new variant along with 73 reference sequences of different clades and variants by MEGA X revealed that strains of the new variant emerged from GR clade (B.1.1) and formed a new cluster (**Figure 1**). In consistent with this result, the phylogenetic dendrogram produced by Nextclade^bera^ v.0.14.3 also showed a new cluster that encompasses the 70 sequences of the new variant within the 20B clade (GR clade) (**Figure 2**). We propose to name this variant as GRL or lineage B.1.1/S:V1230L or 20B/ S:V1230L due to the presence of V1230L in S glycoprotein along with GR clade specific mutations (**Figure 3**). Interestingly, all the 70 sequences of this new variant have been exclusively observed in the state of West Bengal out of 412 sequences (16.99%) submitted from there and distributed in 8 different districts (Kolkata, n=23; Nadia, n=4; North 24 Parganas, n=21; Howrah, n=10; Hooghly, n=4; South 24 Parganas, n=4; Bardhaman, n=2; Paschim Medinipur, n=2) (**Supplementary Table 1**). Among 70 strains, 34, 26 and 10 strains were collected during January, February, and March 2021 respectively. In addition to 12 co-existing mutations including E484K, 113 different mutations were found throughout the genome of these 70 SARS-CoV-2 sequences. Among these 113 mutations, 21 were found in NSP3, 16 were found in S glycoprotein, 13 were found in NSP2, 8 were found in NS3 and N, 7 were found in NS8, 6 were found in NSP6, 5 were found in both NSP5 and NSP16, 4 were found in each of NSP4, NSP12, NSP14 and NS7a, 3 were found in NSP15, 2 were found in E gene, and single mutation was found in each of NSP8, NSP9 and NS6. No mutation was observed in NSP1, NSP7, NSP10, NSP11, NSP13, M and NS7b genes (**Table 2, Supplementary Table 1**).

**Table 1:** Co-existing mutations present within the new variant

| Gene name | Mutation(s) |
| --- | --- |
| NSP4 | D279N, L353F |
| NSP8 | V26F |
| NSP12 | P323L |
| S | D614G, P681H, V1230L, ±E484K |
| NS3 | G172C |
| NS8 | V62L |
| N | R203K, G204R |

**Figure 1:**
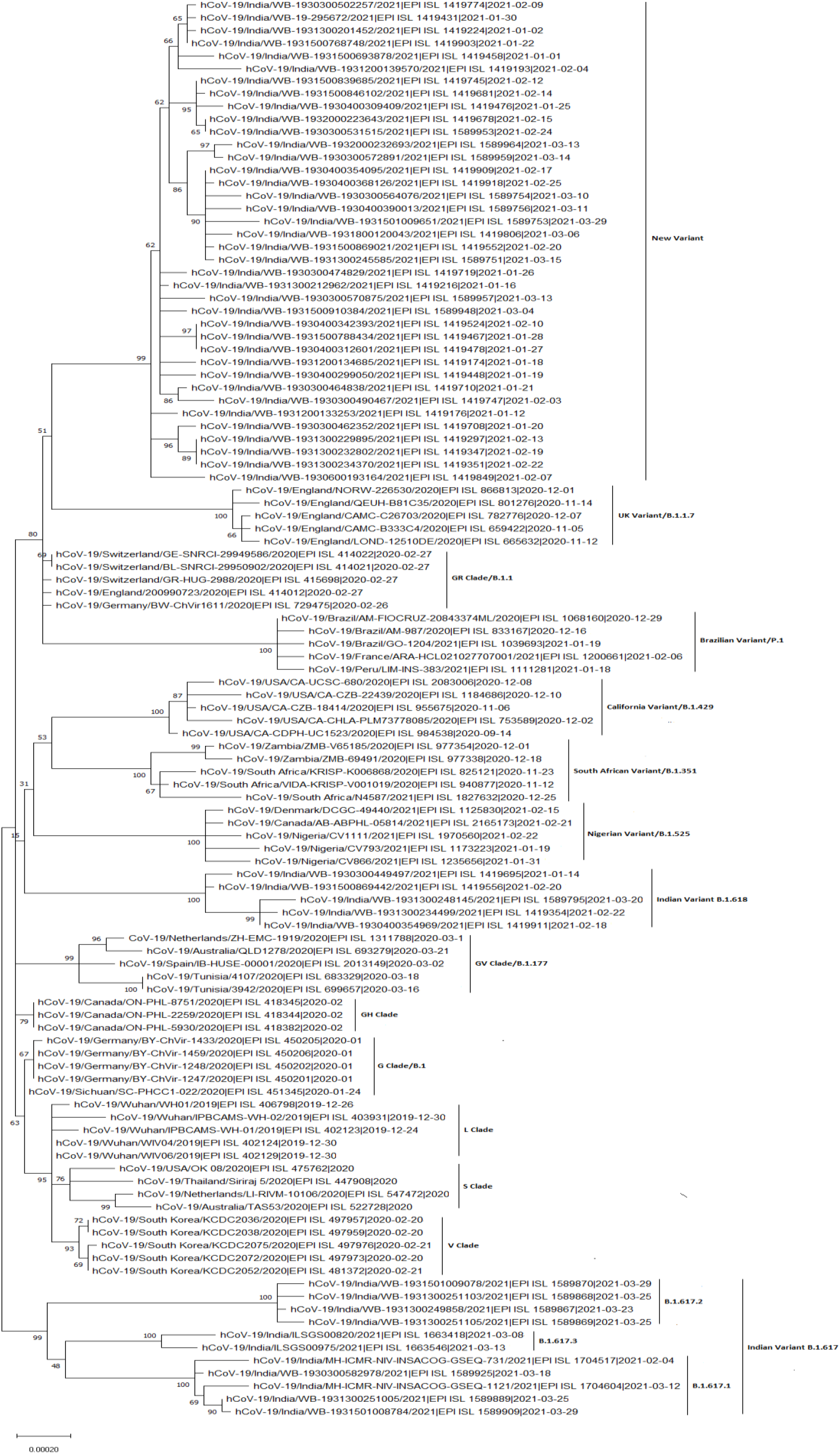
Molecular phylogenetic analysis of new variant strains by MEGA X using maximum-likelihood method. The phylogenetic dendrogram is based on whole genome sequences of 38 representative strains of new variant and 73 reference strains of 16 clades/variants. The scale bar represents 0.00020 nucleotide substitution per site. The best-fit model used for constructing the phylogenetic dendrogram was the general time-reversal model (GTR).

**Figure 2:**
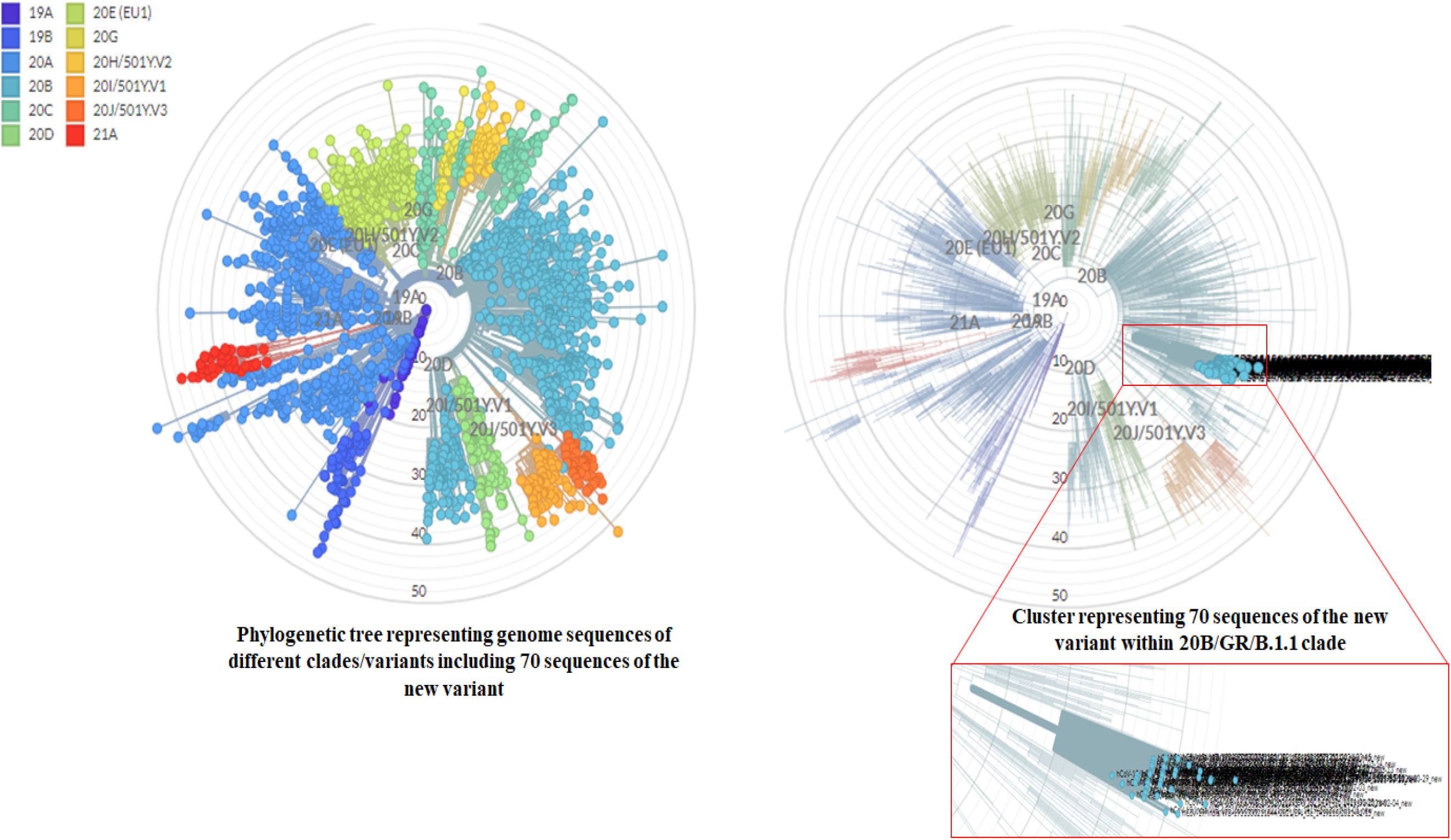
Molecular phylogenetic analysis of new variant strains by Nextclade^beta^ v.0.14.3. Left phylogenetic tree representing all the reference strains of different Nextstrain clades (Automatically selected by Nextclade) as well as 70 strains of the new variant. Each color depicting a different clade. Right phylogenetic tree highlighting only the 70 sequences of the new variant.

**Figure 3:**
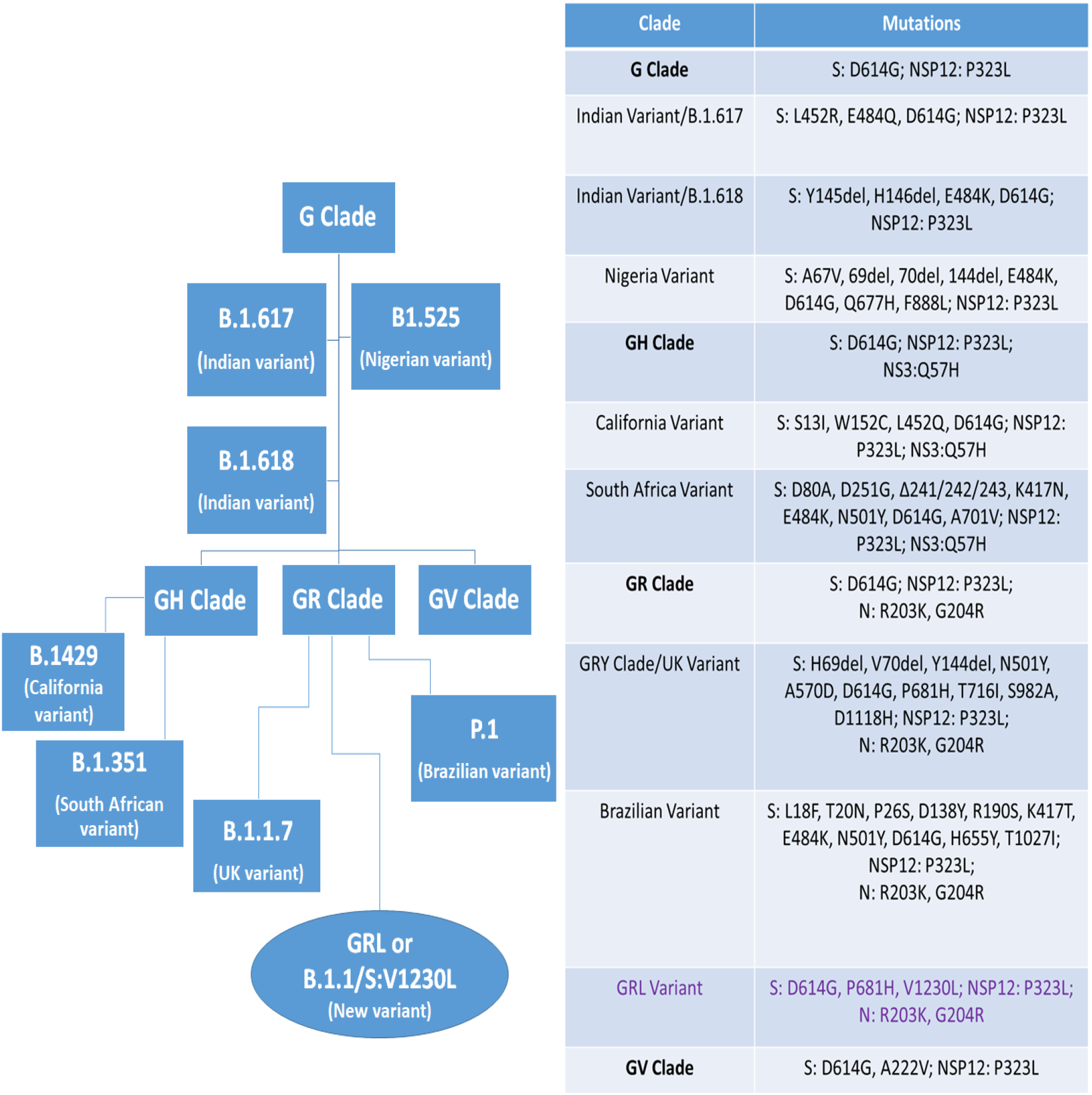
A schematic presentation illustrating the evolution of various clades and variants with their characteristic signature mutations.

**Table 2:** Mutations, excluding the co-existing mutations, found in the different genes of 70 SARS-CoV-2 sequences of the new variant.

|  |  |  |  |  |  |  |  |
| --- | --- | --- | --- | --- | --- | --- | --- |
| **NSP2** |  | **NSP4** |  | A274S | 1 | G251C | 1 |
| **Mutation** | **Frequency** | **Mutation** | **Frequency** | **NSP15** |  | P262S | 1 |
| V94L | 1 | H31Y | 1 | **Mutation** | **Frequency** | D265Y | 1 |
| P129S | 1 | V210I | 1 | S261L | 1 | **E** |  |
| Q134K | 1 | L264F | 1 | S288Y | 1 | **Mutation** | **Frequency** |
| T153M | 1 | T295I | 1 | D300N | 1 | T9I | 1 |
| R222C | 2 | **NSP5** |  | **NSP16** |  | S60T | 1 |
| I224T | 1 | **Mutation** | **Frequency** | **Mutation** | **Frequency** | **NS6** |  |
| I273T | 2 | L58F | 1 | A34V | 1 | **Mutation** | **Frequency** |
| A318S | 2 | K90R | 2 | L126F | 1 | W27L | 1 |
| S430A | 1 | A129V | 2 | T151I | 1 | **NS7a** |  |
| W450C | 1 | T196M | 6 | K160R | 1 | **Mutation** | **Frequency** |
| L451F | 1 | S284G | 1 | 1 | 1 | 1 | 1 |
| V480D | 1 | **NSP6** |  | **S glycoprotein** |  | T39I | 1 |
| V594F | 1 | **Mutation** | **Frequency** | **Mutation** | **Frequency** | P84L | 1 |
| **NSP3** |  | L37F | 1 |  | L5F | Q90R | 1 |
| **Mutation** | **Frequency** | S106F | 1 | P26S | 2 | **NS8** |  |
| D135Y | 1 | K109N | 6 | Q173H | 2 | **Mutation** | **Frequency** |
| A150V | 1 | S118L | 3 | G184V | 2 | T11I | 6 |
| M196I | 1 | V149F | 6 | W258S | 2 | Q18stop | 11 |
| P340L | 1 | G277S | 2 | V382L | 1 | P36S | 1 |
| V477F | 1 | **NSP8** |  | V622F | 1 | R52I | 1 |
| K525R | 2 | **Mutation** | **Frequency** | T681A | 1 | S67F | 3 |
| S609I | 1 | P133S | 1 | A845S | 1 | L95F | 1 |
| A614V | 1 | **NSP9** |  | Q913H | 2 | C102F | 1 |
| T724I | 1 | **Mutation** | **Frequency** | V952F | 1 | **N** |  |
| P822S | 7 | P71S | 1 | V1104I | 2 | **Mutation** | **Frequency** |
| M829V | 1 | **NSP12** |  | I1130M | 1 | G30R | 2 |
| A861S | 1 | **Mutation** | **Frequency** | K1181N | 1 | D128Y | 1 |
| T936I | 1 | L49I | 1 | K1191N | 1 | G215C | 1 |
| P1044S | 1 | K91R | 1 | L1200F | 2 | P279L | 2 |
| T1184M | 1 | V435I | 1 | **NSP3** |  | Q289H | 1 |
| T1189I | 3 | E919D | 1 | **Mutation** | **Frequency** | P383S | 1 |
| L1259F | 2 | **NSP14** |  | G11R | 1 | D401Y | 1 |
| T1335I | 6 | **Mutation** | **Frequency** | V112I | 1 | Q418H | 1 |
| T1379I | 3 | V125F | 1 | C133F | 1 | **Total**<br>**number of**<br>**different**<br>**mutations** | **113** |
| C1392F | 1 | T131I | 1 | V163L | 1 |  |  |
| A1711V | 1 | L152I | 2 | D201Y | 1 |  |  |

Emergence of new mutations in the S glycoprotein, which is involved in the binding of SARS-CoV-2 to the host cell receptor angiotensin-converting enzyme 2 (ACE2) and entry into the host cell, could have influence on virus binding, entry, immune invasion and antibody neutralization. The D614G, the first mutation appeared in the S glycoprotein, was first observed in Germany in January 2020 and became the dominant mutation in all the circulating strains worldwide by June, 2020 [4]. Now it is present in all the circulating strains including the newly emerged VOI (Variant of Interest) and VOC [20]. Patients infected with D614G mutant had higher nasopharyngeal viral RNA loads, indicating its role in increased infectivity [4]. Afterwards, experimental evidences confirmed that the D614G variant enhances virus replication in human lung epithelial cells and primary human airway tissues by increasing the infectivity and stability of the virions. Hamster infected with D614G variant produces higher infectious titers in nasal washes and the trachea, supporting the role of the G614G mutation in high transmissibility. It has also been demonstrated that D614G mutation confers higher susceptibility to serum neutralization [21]. Another study showed that pseudovirus particles carrying S-G614 enter ACE2 overexpressing cells more efficiently than those with S-D614. This increased entry correlates with less S1 domain shedding and higher S protein incorporation into the virion. However, S614 does not alter S protein binding to ACE2 or neutralization sensitivity to pseudoviruses [22]. P681 is located adjacent to the polybasic furin cleavage site _681_P-R-R-A-R-S_686_ where furin mediated proteolytic cleavage is expected to occur between arginine (R_685_) and serine (S_686_). This furin cleavage site is positioned at the junction between S1 domain, essential for virus binding to ACE2 receptor, and S2 domain, necessary for the fusion between virus and the cell membrane. The furin mediated cleavage at the junction of S1/S2 is essential for virus entry into the host cells [23-24]. Therefore, any mutation at this site could influence S1/S2 cleavability by furin-like proteases and hence the infection properties of the virus. The P681H mutation has previously described in the UK variant [7]. A recent study has shown that P681H mutation increases spike protein cleavage by furin-like proteases but does not efficiently impact viral entry and cell-cell spread [25]. The E484K mutation, present within the receptor binding domain of S glycoprotein, has previously reported in Brazilian variant (P.1) and South African variant (B.1.351) [8, 10]. SARS-COV2 variants having E484K mutation is less sensitive to neutralization by serum antibodies induced by previous infection or vaccination [26]. The V1230L mutation, not observed previously in other country, is located at the transmembrane (TM) domain of S2 subunit of S glycoprotein. This transmembrane domain anchors S glycoprotein to the virus envelope. The replacement of valine with more hydrophobic amino acid leucin could tighten the association of S glycoprotein to the viral envelope and also increase S glycoprotein incorporation into the viral envelope. However, further studies are required to reveal the functional relevance of this mutation.

The new variant having D614G, P681H, V1230L and E484K mutations in the S glycoprotein is expected to have increased infectivity, high transmissibility, and lower sensitivity to neutralization antibodies. As India is already reporting high infection rates in the 2^nd^ wave compared to the 1^st^ wave in 2020, the emerging variant needs to be monitored closely. Therefore, further studies involving full genome sequencing of both severe and moderate COVID-19 cases in this region as well as antibody neutralization assay of this variant are required.

## Data Availability

Data are available on request

## Acknowledgement

We would like to acknowledge the scientists, researchers and laboratory staffs in India for their valued contribution in SARS-CoV-2 genome sequencing and deposition in GISAID. We would also like to applaud GISAID consortium for allowing us the open access to the deposited SARS-CoV-2 sequences.

